# Heterogeneity and Superspreading Effect on Herd Immunity

**DOI:** 10.1101/2020.09.06.20189290

**Authors:** Yaron Oz, Ittai Rubinstein, Muli Safra

**Affiliations:** Raymond and Beverly Sackler School of Physics and Astronomy, Tel-Aviv University, Tel-Aviv 69978, Israel; Blavatnik School of Computer Science, Tel-Aviv University, Tel-Aviv 69978, Israel

## Abstract

We model and calculate the fraction of infected population necessary for herd immunity to occur, taking into account the heterogeneity in infectiousness and susceptibility, as well as the correlation between the two parameters. We show that these cause the reproduction number to decrease with progression, and consequently have a drastic effect on the estimate of the necessary percentage of the population that has to contract the disease for herd immunity to be reached. We discuss the implications to COVID-19 and other pandemics.

## I. INTRODUCTION

The COVID-19 pandemic has had a dramatic impact on the world in recent months, setting in motion a huge effort in diverse research disciplines in an attempt to understand the nature of the disease and the dynamics of the virus’s spread. A question of utmost importance in this context is how many infected individuals it takes to reach herd immunity, where by herd immunity one means that the virus is unable to find enough susceptible hosts to continue its spread and consequently the disease fades out.

Herd immunity is typically expected to be reached when a large fraction of the population becomes immune to the disease and the effective reproduction number (which quantifies the spread) drops below one. The standard estimate for the necessary fraction for herd immunity to be reached is about 60% of the susceptible individuals. This estimate, however, assumes a homogeneous structure of the epidemic spread network, where both the infectiousness and susceptibility of individuals are assumed to be homogeneously distributed.

It is well recognized, however, that the epidemic spread network is not homogeneous but rather heterogeneous, with distinct people being *infectious* (likely to infect others) and *susceptible* (likely to become infected themselves) to different degrees (for reviews see e.g. [1, 2]). The class of individuals with very high secondary infection rates are referred to as superspreaders [3]. An estimate for the COVID-19 pandemic [4] asserts that between 5% to 10% of the infected individuals cause 80% of the secondary infections.

The reasons that different people are infectious and susceptible to different degrees may be, for instance, increased contact with others that can increase both parameters, or hygiene and better protective equipment that can decrease both. Thus, infectiousness and susceptibility are potentially highly correlated. A correlation between infectiousness and susceptibility can significantly affect our estimate for the percentage of the population that must contract the disease for herd immunity to be reached.

If more infectious people are also more susceptible, then our initial estimates of the basic reproduction number (the mean value of secondary infections caused by an infected individual) will be biased leading us to believe that it is much larger than it really is—we are oversampling the infectiousness of more susceptible people. Deviations in the susceptibility can lead to us seeing an early spike in the number of cases as the susceptible are infected, with a sudden drop later on as the disease spreads to less susceptible populations. Furthermore, if more infectious people are also more susceptible, then they will also be infected and develop natural immunity much sooner.

Our aim in this work is to model and calculate the fraction of infected in the population that gives rise to herd immunity while taking into account the heterogeneity in infectiousness and susceptibility, their correlation and the superspreading effect.

In order to analyze the spread of the disease we assign to each individual *a* a susceptibility parameter *S*(*a*) and an infectiousness parameter *I*(*a*) drawn from some probability distributions. *S*(*a*) and *I*(*a*) quantify how likely *a* is to be infected and infect others, respectively. The probability that *a* will transmit the disease to *b*—should *a* be infected—is *I*(*a*) × *S*(*b*) and this is the effective parameter in our analysis. The product of *I* and *S* scales like the inverse of the susceptible population, where typically *S* scales like its inverse and *I* is independent of it.

We measure the progress of the disease as a function of the number of individuals who contracted it. This is the natural governing parameter when considering questions of the type: will the disease fade out as a function of the fraction of the population that got infected. We begin the evolution process of the disease at step *n* = 0 with a certain number of infected individuals and increase the number of infected by one at each step. The initial number of infected individuals does not influence the analysis and we will take it to be one. Our calculation is done by taking an expectation value over all possible scenarios of infection.

We model the system for any distribution of *I, S*, and present a general formula for the behaviour of heterogeneous diseases. We also consider the special case where each individual is infectious and susceptible to the same degree, that is, where the distributions from which *I* and *S* are drawn are highly correlated. We derive a simple analytical result when the infectiousness and susceptibility parameters are, in particular, chosen from a Gamma distribution with scaling and shape parameters *k* and *θ*, respectively—a distribution previously attributed to the infectiousness of COVID-2 [3].

Consider first the general case. Define the average conditional infectiousness *φ*(*s*):

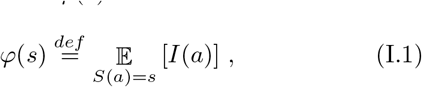

and the normalized susceptibility distribution at the step *n* in the evolution of the disease *ρ*(*s, n*):

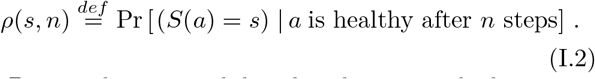

Denote the susceptibility distribution at the beginning by *ρ*(*s*) = *ρ*(*s*, 0). The average conditional infectiousness (I.1) is independent of the number of infected individuals while the susceptibility distribution does depend on it— individuals with higher susceptibility are more likely to be chosen first, hence their rate decreases as the process progresses. We will prove the following general claim about the fraction of the population necessary to reach herd immunity:

**Claim I** (General Case): *for any δ when*

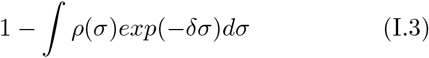

*fraction of the population is infected, the effective reproduction number will be reduced by a factor of*

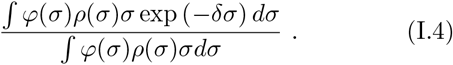

The threshold for herd immunity is when the value of the effective reproduction number is 1.

Consider next the particular case of the Gamma distribution with shape and scale parameters *k* and *θ*, respectively. We will prove the following claim:

**Claim II** (Gamma distribution): *Under the above assumptions, herd immunity will be reached when*

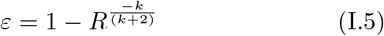

*fraction of the population is infected. R is the reproduction number at the beginning of the disease spread and k is the shape (spread) parameter of the Gamma distribution*.

Note, that the necessary fraction of the population (I.5) does not depend on the scale parameter of the distribution *θ*. Substituting the estimates for COVID-19: *R* ≈ 3 and *k* ≈ 0.1 [5, 6], we get *ε* ≈ 5%. This result is far more optimistic than the recent estimate [7] that requires about 40% of the population to contract the disease before herd immunity is achieved. It also suggests a possible explanation for the observation that the COVID-19 pandemic appears to be slowing down despite the relatively low number of infections so far [4]. The Gamma distribution has been used in order to model the data of COVID-2 with *k* = 0.19 [3] and in this case we get *ε* ≈ 9%.

The letter is organized as follows. In section II we will define precisely the effective reproduction number that we will use in our analysis. In section III we will study the dynamics of the disease spread and prove claim I. In section IV we will prove claim II and study numerically claim I for various types of infection and susceptibility distributions. Section V is devoted to a discussion.

## II. EFFECTIVE REPRODUCTION NUMBER

Consider the effective value of the reproduction number at step *n* in the evolution of the pandemic which we will denote by *R*(*n*), *R* = *R*(0). We define it as the expectation value of secondary infections conditional on the individuals that have been infected. Denote by Λ_*n*_ the distribution over the *n*th individual to be infected (namely, linear in susceptibility at that stage)—we thus have:

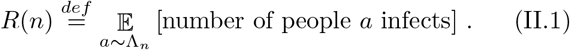

Mathematically we have:

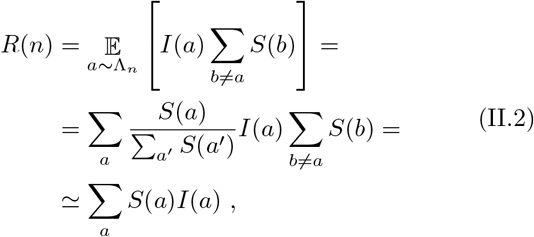

where the summation over *a* in the last two lines is on the healthy individuals at step *n* − 1 and in the approximation at the last line we added the infected individual *a* to the summation. Note, that since *I × S* scales like the inverse of the susceptible population at the *n*th step *N* (*n*), *R* (II.2) does not scale with it. Using (I.1) and (I.2) we get:

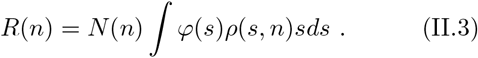

We denote the susceptible population at the beginning by *N* = *N* (0).

In the next section we will study the dynamics of the spread of the disease and how the effective reproduction number *R*(*n*) depends on the number of infected individual.

## III. DISEASE SPREAD DYNAMICS

The process starts at step *n* = 0 with one infected individual. At each step that someone is infected, the probability that *a* was infected is proportional to *S*(*a*). Thus,

Pr [*a* is healthy at step *n*] =

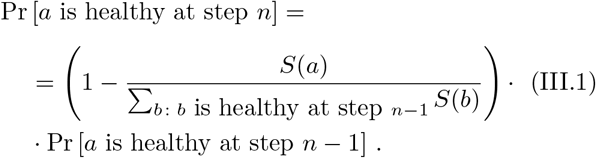

We therefore get:

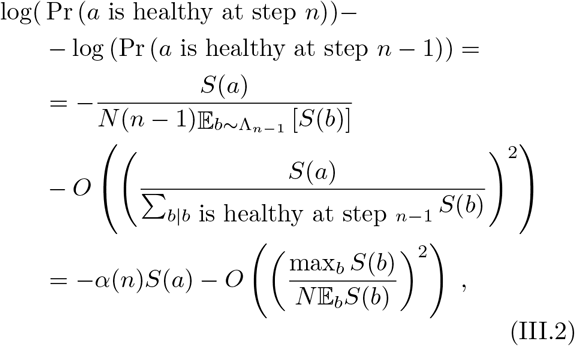

Where

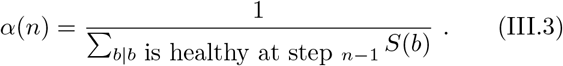

Since at each step someone is infected, there can be at most *N* steps:

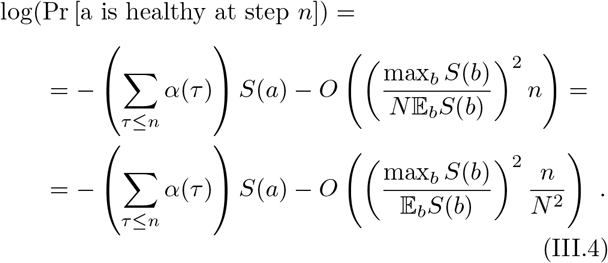

Therefore, as long as 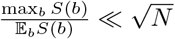 we have:

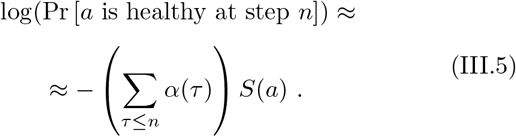

Denote 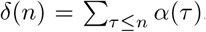, we get a relation between the susceptibility distributions at steps *n* and zero:

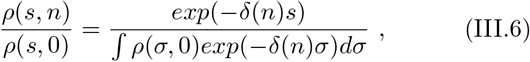

between the susceptible populations:

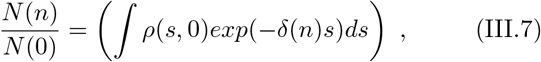

and between the effective reproduction numbers (II.3):

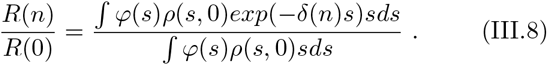

This proves Claim I with (I.3) being 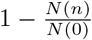.

We can derive the condition for reaching herd immunity at step *n_herd_* by:

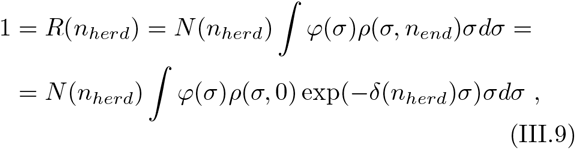

where *N* (*n_herd_*) is related to *N* (*n* = 0) by (III.7).

## IV. HERD IMMUNITY

Consider first the particular case where the infectiousness and susceptibility *I ~ S* and both are drawn from a Gamma distribution:

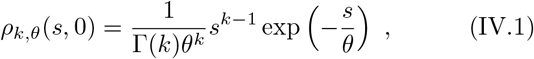

where *k* and *θ* are shape and scale parameters of the distribution at *n* = 0. Using (III.6) we get:

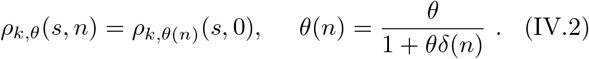

Thus, while the shape of the distribution does not change during the evolution of the disease its scale does.

We denote 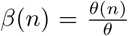, then using (III.7) and (III.8) we get:

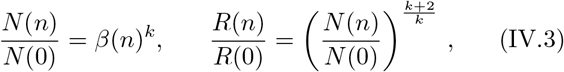

where

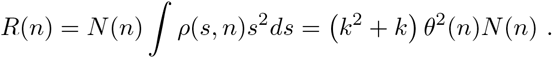

Herd immunity is reached when *R*(*n*) drops below one and this happens when

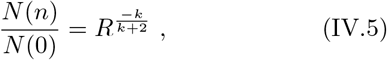

thus proving claim II.

Calculating the fraction of the population *ε* leading to herd immunity, following claim I, can be carried out analytically for the case of a Gamma distribution, while for general distributions it has do be performed numerically. In figures 1 and 2 we plot *ε* as a function of the coefficient of variation, i.e. the ratio of the standard deviation and the mean, 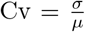. We plot the results for three distributions: Gamma, Folded Normal (Truncated Gaussian) and Power Law when *R* equals three and six, respectively.

**FIG. 1:**
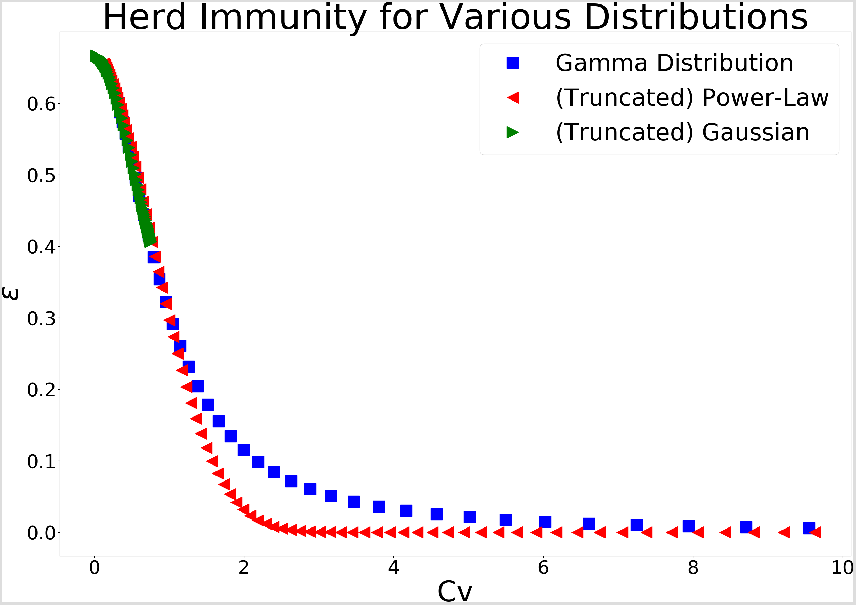
The percentage of infected in the population necessary for herd immunity to occur as a function of the coefficient of variation for *R* = 3. The results are shown for three distributions: Gamma, Folded Normal and Power Law. The higher the variance of the infectiousness and susceptibility the lower the fraction of the population necessary.

**FIG. 2:**
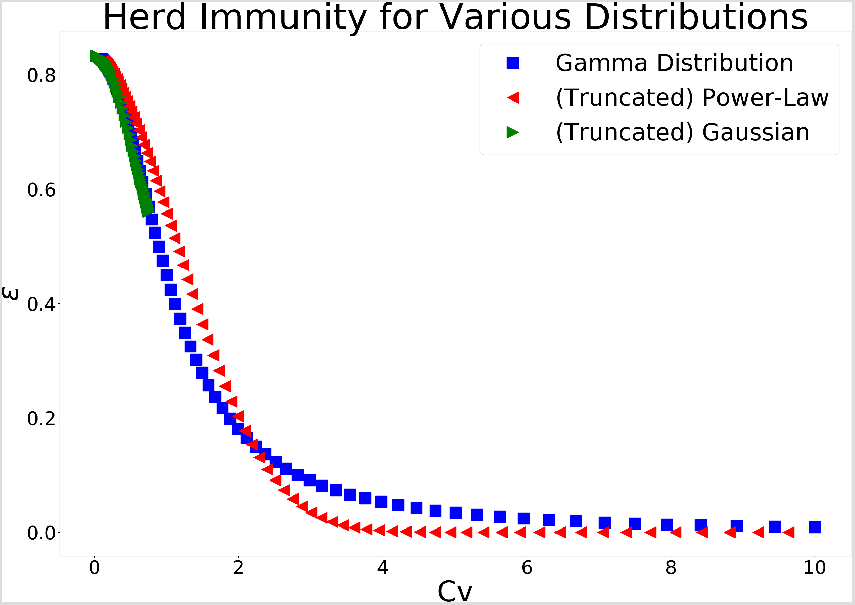
The percentage of infected in the population necessary for herd immunity to occur as a function of the coefficient of variation for *R* = 6. The results are shown for three distributions: Gamma, Folded Normal and Power Law. In comparison to figure 1 we see, as expected, that the required fraction is higher for a given coefficient of variation.

We set *φ*(*s*) = *s*, hence *R* is the second moment of the distribution. We can see that as Cv approaches 0, the distributions behave similarly. However, for larger values, the behaviour of the system depends upon the distribution with the power law distribution approaching *ε* = 0 much faster. The coefficient of variation of truncated Gaussian distribution converges to 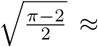 0.755, that is its value for the Half-Normal distribution to which the distribution converges as 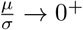.

In general, we see that the higher the variance of the infectiousness and susceptibility the lower the fraction of the population that needs to be infected in order to reach herd immunity.

## V. DISCUSSION

The spread of the COVID-19 pandemics is characterized by high variance of the infection and susceptible distributions. In addition to the high degree of heterogeneity in infectiousness and susceptibility, one expects a significant correlation between them stemming, for instance, from the social aspect of the spread of diseases. We studied the implications of this structure on the condition to reach herd immunity.

We proved two claims, one for general distributions and one for the Gamma distribution and showed that the heterogeneity and correlation have a drastic effect on the estimate of the percentage of the population that must contract the disease before herd immunity is reached.

Under the assumption of Gamma distribution we found that for COVID-19 a fraction *ε* ≈ 5% of infected population suffices to reach herd immunity while a fraction *ε* ≈ 9% is needed for COVID-2. While writing the paper we became aware of two recent works [9, 10] that, using different mathematical fraemeworks, reached similar conclusions for the fraction of infected population that is required for herd immunity under the assumption of Gamma distribution.

Our mathematical analysis of the disease spread dynamics can be viewed as a random walk on a complete graph. It is of interest to study other graph structures and quantify the differences [11].

## Data Availability

all data used in the paper is available on line or in scientic papers cited

## Acknowledgements

We would like to thank Nir Kalkstein for valuable discussions on the importance of the high variance to the spread of the disease. The work is supported in part by the Israeli Science Foundation center of excellence, the European Research Council (ERC) under the European Union’s Horizon 2020 research and innovation program (Grant agreement No. 835152), as well as by ISF 2013/17 and BSF 2016414.

